# Barriers and Facilitators to Heart Failure Guideline-Directed Medical Therapy in an Integrated Health System and Federally-Qualified Health Centers: A Thematic Qualitative Analysis

**DOI:** 10.1101/2024.10.28.24316301

**Authors:** Sarah E. Philbin, Lacey P. Gleason, Stephen D. Persell, Eve Walter, Lucia C. Petito, Anjan Tibrewala, Clyde W. Yancy, Rinad S. Beidas, Jane E. Wilcox, R. Kannan Mutharasan, Donald Lloyd-Jones, Matthew J O’Brien, Abel N. Kho, Megan C. McHugh, Justin D. Smith, Faraz S. Ahmad

## Abstract

**Background:** Clinical guidelines recommend medications from four drug classes, collectively referred to as quadruple therapy, to improve outcomes for patients with heart failure with reduced ejection fraction (HFrEF). Wide gaps in uptake of these therapies persist across a range of settings. In this qualitative study, we identified determinants (i.e., barriers and facilitators of quadruple therapy intensification, defined as prescribing a new class or increasing the dose of a currently prescribed medication.

**Methods:** We conducted interviews with physicians, nurse practitioners, physician assistants, and pharmacists working in primary care or cardiology settings in an integrated health system or Federally Qualified Health Centers (FQHCs). We report results with a conceptual model integrating two frameworks: 1) the Theory of Planned Behavior (TPB), which explains how personal attitudes, perception of others’ attitudes, and perceived behavioral control influence intentions and behaviors; and 2) The Consolidated Framework for Implementation Research (CFIR) 2.0 to understand how multi-level factors influence attitudes toward and intention to use quadruple therapy.

**Results:** Thirty-one clinicians, including thirteen eighteen (58%) primary care and (42%) cardiology clinicians, participated in the interviews. Eight (26%) participants were from FQHCs. A common facilitator in both settings was the belief in the importance of quadruple therapy. Common barriers included challenges presented by patient frailty, clinical inertia, and time constraints. In FQHCs, primary care comfort and ownership enhanced the intensification of quadruple therapy while limited access to and communication with cardiology specialists presented a barrier. Results are presented using a combined TPB-CFIR framework to help illustrate the potential impact of contextual factors on individual-level behaviors.

**Conclusions:** Determinants of quadruple therapy intensification vary by clinician specialty and care setting. Future research should explore implementation strategies that address these determinants by specialty and setting to promote health equity.

## Background

Heart failure (HF) is a condition with high morbidity and mortality that affects the physical and mental of over 6.5 million US adults with a disproportionate impact on populations experiencing disadvantage.^1,2^ For the approximately 40% of the HF population with HFrEF, randomized controlled trials have demonstrated the profound impact of medications from four drug classes at target dosing.^3^ Collectively referred to as foundational GDMT or quadruple therapy, these medications improve quality of life, reduce the risk of hospitalization and mortality by 64%, and add an estimated eight life-years with comprehensive treatment with all 4 drug classes.^4,5^ Yet, data from US registries and health systems show consistently suboptimal use of these life-saving therapies.^6–12^

Determinants, or barriers and facilitators, of quadruple therapy intensification, defined as adding a new class of medication or increasing the dose on an already prescribed medication, have been examined using quantitative and qualitative approaches in primary care and cardiology practices in large health systems.^10,13–17^ FQHCs are one of the main sites of primary care delivery in the U.S., especially for patients, who are underinsured, uninsured, or have Medicaid. However, determinants of HF care in FQHCs have not been examined, although patients seen in this care setting are predominately from groups that are low-income, have the highest morbidity and mortality from HF, and have limited specialty care access^18^. Due to the lack of scaling of previously tested strategies to increase GDMT prescription, better characterizing determinants across diverse settings will facilitate tailored implementation strategies by care setting and may contribute to increasing equitable outcomes for all patients with HF.

In this study, we sought to identify determinants of prescribing quadruple therapy from different specialties (primary care and cardiology) and care settings (large integrated health system and FQHCs) with a previously-investigated conceptual model^19,20^ that integrates two, widely-used theories and frameworks from the social science and implementation science literature: the TheoryPB^21^ and the CFIR 2.0.^22^

## Methods

### Study Design and Recruitment Procedures

We conducted a qualitative study with semi-structured interviews of clinicians. The study followed the COREQ Checklist for reporting (Supplemental Table 1).^23^ The Northwestern University Institutional Review Board approved this study. All participants provided verbal informed consent.

Participants were recruited from a multi-county integrated health system and a multi-state FQHC network. Eligible participants included physicians, nurse practitioners, physician assistants, and pharmacists in cardiology and primary care. We used a purposive sampling approach.^24^ Details on recruitment procedures is provided in the Supplement.

### Data Collection

Data were collected during one-on-one, 30-minute virtual interviews. The interviews were conducted using semi-structured interview guides tailored to clinician specialty. Using CFIR, we developed questions to identify factors that impact the implementation of evidence-based care. We developed additional questions based on the Chronic Care Model, because this model includes specific elements of health systems associated with promoting high quality, evidence-based care.^26,27^ The interview guides and additional details on their development are included in the Supplement.

### Analysis and Theoretical Framework

Professionally transcribed interviews were analyzed in ATLAS.ti (Berlin, Germany)^28^ using thematic analysis, which reports themes identified in the data.^29^ We developed a set of codes a priori that were informed by the interview guide and literature. Two coders (LPG and SEP) independently coded 20% of the integrated health system and 20% of the FQHC transcripts and met to review discrepancies and the emergence of new themes from the data. A detailed description of the coding process is included in the Supplement. Although the sample of FQHC participants was smaller than that of the integrated health system, the findings from the FQHC transcripts reached thematic saturation. After coding was complete, FSA, LPG, and SEP independently prepared analytic memos, which allows team members d to synthesize data into high-level themes.^31^ The memos addressed six questions (Supplemental Table 6) and were compared to identify common preliminary themes. In a post-hoc, analytical decision, we elected to report findings in a previously investigated TPB-CFIR framework to better define the relationships between contextual multilevel determinants and clinician intention and action to intensify quadruple therapy.^19,20^ The TPB purports that intention is the most proximal contributor to behavior and is influenced by three constructs. The first construct, attitude, encapsulates the degree of favorability that an individual ascribes to a behavior. The second construct, subjective norms, references the social pressure surrounding behaviors. The third construct, perceived behavioral control, encompasses the perception of the ease or difficulty of performing the behavior. The lack of incorporation of multi-level determinants influencing beliefs and intentions represents a limitation of the TPB.^20^ The integration with CFIR 2.0, which comprehensively covers five multilevel ecological domains influencing implementation outcomes, provides a holistic picture of factors influencing quadruple therapy intensification from the clinician perspective.

## Results

### Participant Demographics

We conducted a total of 31 interviews, including 23 health system and 8 FQHC clinicians (Table 1). The entire sample (n=31) was predominantly female (61%) with a median age of 39 years (Table 2).

**Table 1.**
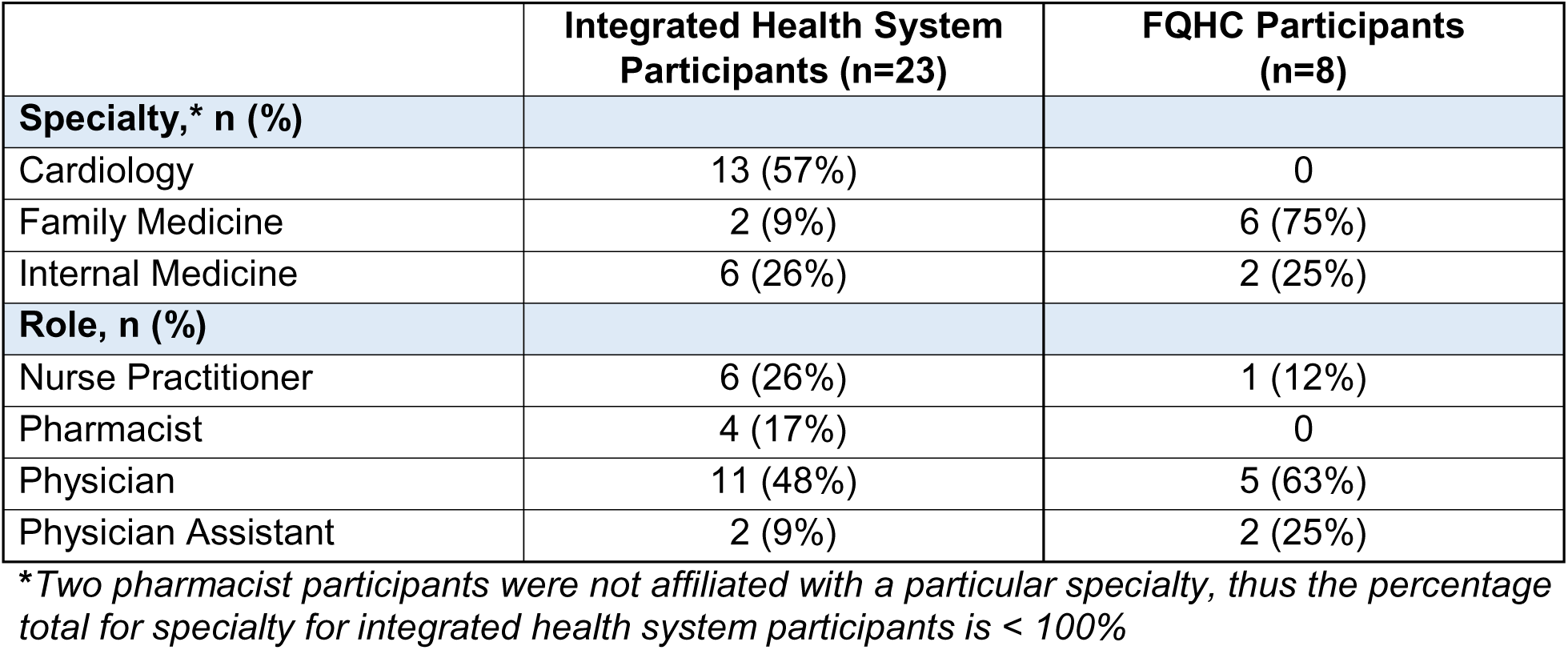
Participant Clinical Specialties and Roles.

**Table 2.** Participant Demographics.

|  |  |
| --- | --- |
| **Age\*, median in years (IQR)** | 39 (17) |
| **Sex, n (%)** |  |
| Female | 19 (61%) |
| Male | 12 (39%) |
| **Ethnicity,\*\* n (%)** |  |
| Not Hispanic or Latino | 31 (100%) |
| Hispanic or Latino | 0 (0) |
| Other | 1 (3%) |
| **Race,\*\* n (%)** |  |
| American Indian or Alaska Native | 0 (0%) |
| Asian | 5 (16%) |
| Black or African American | 1 (3%) |
| Native Hawaiian or Other Pacific Islander | 0 (0%) |
| White | 24 (77%) |
| Other | 2 (7%) |

*\* 2 participants declined to report age and are not included in the mean*
*\*\*Race and Ethnicity were collected as all that apply. Thus, the percentage totals >100%*

### Theory of Planned Behavior Constructs and CFIR Domains

A summary of thematic findings is presented in Figure 1.

**Figure 1.**
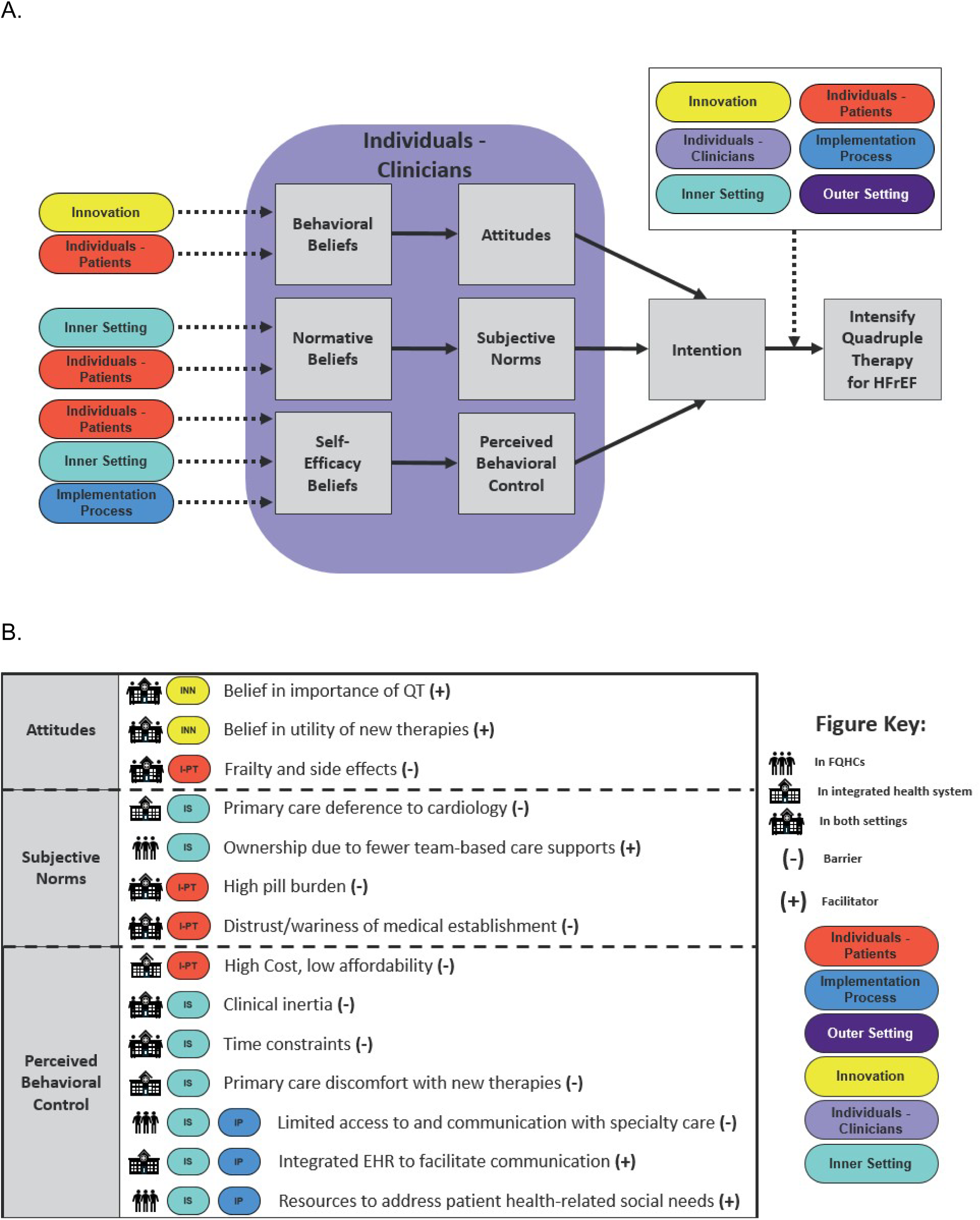
Applications of Theory of Planned Behavior and CFIR to a Cardiology or Primary Care Visit. Figure 1A depicts the relationship of CFIR domains, including innovation, individuals, inner and outer setting, and implementation processes on Theory of Planned Behavior constructs of attitude, subjective norms, and perceived behavioral control. The Theory of Planned Behavior constructs influence clinician intention and subsequent use of quadruple therapy for patients with HFrEF at the time of clinical visits in cardiology and primary care. The Figure 1 Key lists the findings that map to the CFIR domains and TPB constructs. It also provides an overview of symbols that indicate if a determinant is a barrier or facilitator and if it is specific to the FQHC setting, integrated health system setting, or both. EHR=Electronic Health Record; FQHC = Federally Qualified Health Center; HFrEF=Heart Faiure with Reduced Ejection Fraction; QT = quadruple therapy

#### CFIR Domains that Inform Attitudes

##### Innovation: The “thing” being implemented (Intensification of quadruple therapy)

Participants described their attitude toward the prescription of quadruple therapy. As illustrated through the quote in Table 3, there was unanimous support for the importance of quadruple therapy. Many participants’ perspective on their role in the prescription of quadruple therapy was influenced by their knowledge of the evidence base for quadruple therapy. Participants expressed that changes to the evidence base, such as the addition of new medications to patients’ regimens, was an important component of treatment. For example, participants noted that some of the medication classes, including ACE Inhibitors, ARBs, and beta blockers, have served as the longstanding foundation of HFrEF treatment. Some participants noted that when SGLT2i were added to the guidelines, they were less familiar with the medication class but have become more familiar and recognize the value of this medication class.

**Table 3:** Illustration of TPB and CFIR Constructs through Participant Quotations.

| TPB Construct and CFIR Domain | Quotation |
| --- | --- |
| **TPB Construct: Attitudes**<br>**CFIR Domain: Innovation** | *It's important because it helps them [patients] hopefully get a little bit better outcomes and helps them hopefully stay out of the hospital. Helps them feel better. Symptom control. Hopefully prolonging their life or helping them potentially with quality of life.* |
| **TPB Construct: Subjective Norms**<br>**CFIR Domain: Inner Setting** | *I have had patients that I've considered prescribing them something like ARBs, but I would say, 99 percent of the time, what I would do is, if the patient has a cardiologist, send them a message of saying, "Hey. What do you think about an ARNI in this patient?" Then leave it up to them to decide whether or not to prescribe it.* |
| **TPB Construct: Perceived Behavioral Control**<br>**CFIR Domain: Inner Setting** | *With heart failure physicians this is what your primary job is. What you're in-tuned to doing or attuned to doing every day. But when I get referrals for new patients, I would say that clinician inertia is a big part of the problem for sure. [...] You'll still see people who would qualify for [titration] who are left on an ACE inhibitor ARB with no reason to have not tried switching over. People who might—not even a—maybe it's inertia. Because the patient, "feels okay", but will be on a non-guideline directed beta blocker—like atenolol, for example, for hypertension which carry over when they developed heart failure. That should be switched over to one of the indicated beta blockers for HFrEF.* |
| **TPB Construct: Perceived Behavioral Control**<br>**CFIR Domain: Individuals - Patients** | *I think the cost surrounding these agents and insurance coverage. We never know, especially for Medicare patients with all the different supplemental plans. I think it's very difficult to know what the coverage is. Especially for ARBs and the SGL2 inhibitors. Usually we end up sending it to the pharmacy. Then, calling the pharmacy and doing a price check. I think if there were some kind of resource. I don't know if this'll ever be possible. If we could price check it* |
|  | *before sending it over. That we could tell the patient what they could expect in terms of cost. Would be super helpful. I think that's maybe sometimes what limits inpatient initiation of those agents, too. The physicians don't wanna send the patients home with a huge cost worth of medications. It's hard to tell what the cost is gonna be or what insurance is gonna cover* |
| **TPB Construct: Perceived Behavioral Control**<br>**CFIR Domain: Implementation Process** | *Unfortunately, I think that's a fragmented issue at least in our healthcare system because we are not necessarily affiliated with any particular tertiary hospital center. A lot of times, it requires me to be on top of the patient's care, especially in terms of—a lot of times I'm unaware of the fact that they went to see the cardiologist or that the cardiologist changed their medications. I would say that probably happens about 75 to 80 percent of the time that I'm unaware of it.* |

##### Individuals - Patients: Patient-related factors that impact quadruple therapy intensification

Patients are the recipients of the innovation, quadruple therapy. There are some recipient characteristics, including need and capability, which impact experience with quadruple therapy. Participants described challenges associated with quadruple therapy-related decision-making for frail patients. Several participants noted that side effects, such as hypotension, that accompany some of the medication classes, may be especially problematic for these patients. In some instances, side effects informed participants’ attitude toward initiating or adjusting quadruple therapy. These side effects represent needs that can limit quadruple therapy intensification.

#### CFIR Domains that Inform Subjective Norms

##### Inner Setting: Factors related to the site that impact quadruple therapy intensification

Participants described the impact of subjective norms and the inner setting of their clinical environment on their decision to prescribe or adjust quadruple therapy. In contrast to cardiology participants and as illustrated by the quote in Table 3, primary care participants reported variable comfort with prescribing and increasing the dose of ARNIs. A clinician’s understanding of the division of responsibility between primary care and cardiology and in the management of different diagnoses may influence their willingness to take ownership of HF management. Among participants, this phenomenon was most apparent in primary care clinicians’ deference to cardiologists regarding the management of HFrEF medications. Even in instances where primary care clinicians noted that they were willing to adjust HF medications, many noted the importance of consulting with the cardiologist prior to making changes. Participants from the integrated health system reflected on a collaborative approach to care that included physicians, advanced care practitioners, and pharmacists. Alternatively, FQHC-based participants observed that this team-based care approach for HFrEF was less common in their practices with more of the responsibility for quadruple therapy management falling to primary care clinicians.

##### Individuals-Patients: Patient-related factors that impact quadruple therapy intensification

When speaking to patient knowledge, many participants noted that their patients do not always understand the importance of quadruple therapy and grow frustrated by pill burden, especially when taking medications for multiple conditions. Another common patient-related need involved addressing wariness of the medical establishment. Some participants shared that their patients expressed concerns about side effects associated with quadruple therapy that they heard in television commercials. Others reflected that their patients were hesitant about the use of quadruple therapy because they preferred nonpharmaceutical remedies, such as dietary changes. While these patient-level determinants present barriers to the prescription of quadruple therapy, many clinician participants highlighted the potential role of patient education as a mechanism to achieve target medication doses. Participants offered examples of patient education initiatives, including HFrEF-related animations and teach back techniques, that can help patients understand the diagnosis and roles of medication classes in symptom management.

#### CFIR Domains that Inform Perceived Behavioral Control

##### Individuals-Patients: Factors related to patients that impact quadruple therapy intensification

The most frequently mentioned patient-related factors impacting perceived behavioral control were cost and affordability. Participants noted that the cost of specific medications, including ARNIs and SGLT2i, may be particularly burdensome to some patients. FQHC-based participants also reflected on the role that patient population characteristics played in their decision to initiate and titrate HF medications. While integrated health system-based participants described the challenges that comorbidities present in the management of HF medications, FQHC-based participants highlighted the challenges presented by specific comorbidities, such as substance use disorders and the impacts of social determinants of health, including housing insecurity. FQHC-based participants more frequently mentioned the impact of patients’ health literacy and fewer touchpoints with the health system due to less frequent appointments.

##### Inner Setting: Factors related to the site of implementation of quadruple therapy

Participants identified several factors in the inner setting that influence behavioral control and may facilitate or hinder the prescription of quadruple therapy. First, some primary care participants described the impact of clinical inertia on the culture of their clinical setting, noting that they may be hesitant to change a medication dose if the current dose was well-tolerated. In contrast, cardiology participants, such as the one quoted in Table 3, described less hesitancy about initiating new medications or up-titrating existing medications when patients are doing well. In fact, they were focused on continuing to make changes to adhere to guidelines. Time constraints may also hinder a clinician’s ability to optimize quadruple therapy for HFrEF and impacts the innovation’s compatibility with the inner setting. Some participants described factors that may aid in surmounting time constraints, including advanced practice clinician-facilitated communication between primary care and cardiology to discuss potential changes to the HF medications. Participants noted that access to effective and reliable communication mechanisms between cardiology and primary care may also impact the prescription of quadruple therapy. While participants in the integrated health system described the benefits of an EHR system that facilitated communication between primary care and cardiology and between patients and clinicians, FQHC-based participants noted the challenges they faced communicating with cardiology specialists, who were outside of their health system.

When the issue of cost and affordability was discussed, there was some indication from FQHC-based participants that existing programs for their patients help overcome these barriers, which speaks to the role that local conditions play in supporting the implementation of quadruple therapy. More specifically, many of their patients are on Medicaid and qualify for income-based patient assistance programs that help to reduce medication costs. Other participants described the role of organizational-level programs, such as the 340B Program, which allow participating organizations to procure outpatient medications at a discount. While these programs offer avenues to address affordability challenges, some participants noted that challenges persist, especially surrounding costs of SGLT2i and ARNIs.

##### Implementation Process: Activities and strategies used to implement quadruple therapy

A comparison of findings from the participants who practice in integrated health systems vs. FQHCs indicates some differences in their experiences with prescribing quadruple therapy. As described above, FQHC-based primary care clinicians often indicated that they were more willing to initiate and titrate HF medications compared to primary care clinicians based at an integrated health system due to inconsistent access to and fragmented communication with cardiology specialists.. Fragmented communication can contribute to a greater burden on the primary care clinicians to determine what changes have been made, and this process can be further complicated by patients’ language barriers. These challenges illustrate the importance of adaptation as a component of the implementation process.

## Discussion

Using the combined TPB and CFIR 2.0 conceptual model, we highlight multi-level determinants that inform the action of quadruple therapy intensification and illustrate how these determinants influence the action of quadruple therapy intensification during clinic visits. We also identified differences between clinician types (i.e., primary care clinicians, cardiology clinicians, pharmacists) and between those from different types of organizations (i.e., integrated health system, FQHCs) regarding their comfort initiating and titrating HF medications. Primary care clinicians were more likely to describe clinical inertia as a barrier to quadruple therapy intensification compared to cardiology clinicians. Among primary care clinicians, those at FQHCs were typically more comfortable managing quadruple therapy compared to their integrated health system counterparts, who often deferred to cardiology clinicians to manage the regimen. Within the integrated health system, co-management was facilitated through EHR-based communication while primary care clinicians based at FQHCs struggled to co-manage patients with cardiology specialists due to lack of integrated EHR and patients’ lack of established relationships with specialists. Both integrated health system and FQHC clinicians identified patient-related factors that complicate HF medication management, including frailty, side effects, and pill burden. FQHC clinicians also highlighted additional patient-related barriers, including health-related social risks like housing insecurity and complex co-morbidities including substance use and mood disorders.

This study extends the growing literature on barriers and facilitators to prescription of evidence-based therapies in HF.^13–15,32–36^ Our findings are complementary to a taxonomy that accounts for the role of clinical inertia in treatment non-intensification that was previously applied to HFrEF trial data and adds a conceptual model that identifies potential causal mechanisms between multi-level determinants and quadruple therapy intensification.^33^ One qualitative study of cardiology and primary care clinicians at a different integrated health system identified similar challenges related to clinical inertia, patient concerns around pill burden, competing priorities, affordability of SGLT2is and ARNIs, and diffusion of responsibility for care across the health system.^13 16,17^ Our study provides more insight into the role that EHR-based communication between primary care and cardiology clinicians can play in facilitating the management of quadruple therapy and adds a comparison with FQHCs. Our findings are also complementary to those from a study that assessed the perspectives of primary care physicians on their experience co-managing patients with chronic kidney disease with nephrologists and identified barriers related to timely information exchange, unclear roles and responsibilities, and limited access to nephrologists.^37^

Existing research on the implementation of quadruple therapy within FQHCs is limited, but our findings can be compared to the treatment and management of mental health diagnoses within an FQHC setting. Prior qualitative research that assessed clinician perspectives on the treatment of mental health in primary care found that competing priorities, such as patients with multiple health concerns, and discomfort managing psychotropic medications can complicate the management of mental health diagnoses by primary care clinicians.^38^ These results align with our findings from both integrated health system-and FQHC-based primary care clinicians. Other research in this space has demonstrated that FQHC-based clinicians face communication challenges when referring patients to more specialized mental health treatment.^39^ This is similar to findings from our FQHC-based participants, who indicated that they have difficulty referring their patients to cardiology specialists and encounter communication challenges due to lack of integrated EHRs. Finally, interprofessional collaborative care for patients with multiple chronic conditions, including depression and high cholesterol, has been suggested as a mechanism to better serve patients who receive care in FQHCs.^40^ A similar approach might be beneficial for patients with HFrEF in FQHCs given the potential for cross-specialty collaboration in the management of quadruple therapy. Given the documentation of the challenges faced in FQHC settings across different diagnostic categories, it is important to consider these challenges when developing strategies to enhance the use of quadruple therapy for patients with HFrEF.

Quadruple therapy represents an evidence-based practice that can enhance cardiovascular health equity by improving patient outcomes across diverse populations.^36^ Although several implementation strategies have been tested to improve the uptake of quadruple therapy,^41,42^ none, to our knowledge, has prioritized reaching patients from under-resourced communities who primarily receive care in FQHCs. Moreover, implementation studies for quadruple therapy have prioritized the evaluation of process and clinical outcomes, but largely have not evaluated costs or implementation strategy mechanisms.^36,43,44^ Future research should test tailored strategies based on determinants and settings, employ a comprehensive evaluative framework with extensions to promote health equity,^45,46^ and measure costs. A comprehensive evaluation will ultimately increase the likelihood of scaling and sustaining successful implementation strategies.

This study has several limitations and strengths. We had disparate sample sizes from each setting type, which may introduce selection bias. However, this study, to our knowledge, provides the first qualitative assessment of FQHC clinician perspectives on determinants of quadruple therapy for patients with HFrEF, and the analysis of FQHC data did reach thematic saturation. Physicians represented 52% (n=16) of our total sample size. Considering the interdisciplinary nature of care for patients with HF, future research should continue to explore the perspectives of other members of the care team, including pharmacists and advanced care practitioners. Lastly, our study reports clinician-identified, patient-level determinants because interviewing patients from each setting was outside the scope of the goals and resources of the study. Future work will identify barriers and facilitators from the patient perspective across diverse settings, including FQHCs.

## Conclusion

Key barriers to the uptake of quadruple therapy include clinician comfort with medication adjustment, understanding division of responsibility within a collaborative care team, communication challenges between clinicians from different specialties, side effects, and comorbidities. The need to identify implementation strategies to improve the uptake of quadruple therapy that address barriers at the patient-, clinician-, and organizational-level persists. Future research should explore these potential strategies while considering unique barriers faced by patient populations who bear a disproportionate burden of poor HF outcomes and receive care outside of health systems and the different perspectives and needs of primary and cardiology clinicians.

## Data Availability

All data produced in the present study are not available due as permission to share the data was not requested as part of the informed consent process.

## Abbreviations

HFrEF: Heart Failure with reduced ejection fraction
GDMT: Guideline-directed medical therapy
FQHC: Federally-qualified Health Centers
TPB: Theory of Planned Behavior
CFIR 2.0: Consolidated Framework for Implementation Research 2.0
COREQ: Consolidated Criteria for Reporting Qualitative Research
ACE inhibitors: Angiotensin-Converting Enzyme Inhibitors
ARBs: Angiotensin II Receptor Blockers
SGLT2i: Sodium-Glucose Cotransporter-2 inhibitors
ARNIs: Angiotensin receptor/neprilysin inhibitors
EHR: Electronic health record

## Acknowledgements

The content is solely the responsibility of the authors and does not necessarily represent the official views of the National Institutes of Health.

