## Supplemental Material for "Barriers and Facilitators to Heart Failure Guideline-Directed Medical Therapy in an Integrated Health System and Federally-Qualified Health Centers: A Thematic Qualitative Analysis"

**HF GDMT Prescription Determinants**

**Supplemental Material**

**Table of Contents**

### COREQ (Consolidated criteria for Reporting Qualitative research) Checklist – Table 1

| **Item No. and Topic** | **Guide Questions/Description** | | **Reported on Pg. No** | |
| --- | --- | --- | --- | --- |
| **Domain 1: Research team and reflexivity** | | | | |
| *Personal Characteristics* | | | | |
| 1.Interviewer/Facilitator | | Which author/s conducted the interview or focus group? | | Supplement (pg. 5) |
| 2. Credentials | | What were the researcher’s credentials? E.g. PhD, MD | | Cover Page |
| 3. Occupation | | What was their occupation at the time of the study? | | N/A |
| 4. Gender | | Was the researcher male or female? | | N/A |
| 5. Experience and Training | | What experience or training did the researcher have? | | Supplement (pg. 5) |
| *Relationship with Participants* | | | | |
| 6. Relationship Established | | Was a relationship established prior to study commencement? | | Supplement (pg. 5) |
| 7. Participant Knowledge of the interviewer | | What did the participants know about the researcher? e.g. personal goals, reasons for doing the research | | N/A |
| 8. Interviewer Characteristics | | What characteristics were reported about the inter viewer/facilitator? e.g. Bias, assumptions, reasons and interests in the research topic | | N/A |
| **Domain 2: study design** | | | | |
| *Theoretical framework* | | | | |
| 9. Methodological orientation and Theory | | What methodological orientation was stated to underpin the study? e.g. grounded theory, discourse analysis, ethnography, phenomenology, content analysis | | 7-8 |
| *Participant selection* | |  | |  |
| 10. Sampling | | How were participants selected? e.g. purposive, convenience, consecutive, snowball | | Manuscript (pg. 5) and Supplement (pg. 5) |
| 11. Method of approach | | How were participants approached? e.g. face-to-face, telephone, mail, email | | Supplement (pg. 5) |
| 12. Sample size | | How many participants were in the study? | | Manuscript Pg. 9 and Tables 1 and 2 |
| 13. Non-participation | | How many people refused to participate or dropped out? Reasons? | | N/A |
| *Setting* | |  | |  |
| 14. Setting of data collection | | Where was the data collected? e.g. home, clinic, workplace | | Manuscript (pg. 6) |
| 15. Presence of non-participants | | Was anyone else present besides the participants and researchers? | | Manuscript (pg. 6) |
| 16. Description of sample | | What are the important characteristics of the sample? e.g. demographic data, date | | Manuscript (pg. 8, Tables 1 and 2) |
| *Data collection* | |  | |  |
| 17. Interview guide | | Were questions, prompts, guides provided by the authors? Was it pilot tested? | | Supplemental (Tables 2, 3, 4, 5) |
| 18. Repeat interviews | | Were repeat interviews carried out? If yes, how many? | | N/A |
| 19. Audio/visual recording | | Did the research use audio or visual recording to collect the data? | | Supplement (pg. 5) |
| 20. Field notes | | Were ﬁeld notes made during and/or after the interview or focus group? | | N/A |
| 21. Duration | | What was the duration of the interviews or focus group? | | Manuscript (pg. 6) |
| 22. Data saturation | | Was data saturation discussed? | | Manuscript (pg. 7) |
| 23. Transcripts returned | | Were transcripts returned to participants for comment and/or correction? | | N/A |
| **Domain 3: analysis and ﬁndings** | | | | |
| *Data analysis* | | | | |
| 24. Number of data coders | | How many data coders coded the data? | | Manuscript (pg. 7) |
| 25. Description of the coding tree | | Did authors provide a description of the coding tree? | | Manuscript (pg. 7) and Supplement (pg. 5) |
| 26. Derivation of themes | | Were themes identiﬁed in advance or derived from the data? | | Manuscript (pg.7-8) and Supplement (pg. 5) |
| 27. Software | | What software, if applicable, was used to manage the data? | | Manuscript (pg. 7) |
| 28. Participant checking | | Did participants provide feedback on the ﬁndings? | | N/A |
| *Reporting* | | | | |
| 29. Quotations presented | | Were participant quotations presented to illustrate the themes/ﬁndings? Was each quotation identiﬁed? e.g. participant number | | Manuscript (Table 3) |
| 30. Data and ﬁndings consistent | | Was there consistency between the data presented and the ﬁndings? | | Manuscript (9-13, Table 2 |
| 31. Clarity of major themes | | Were major themes clearly presented in the ﬁndings? | | Manuscript (pgs. 9-14) |
| 32. Clarity of minor themes | | Is there a description of diverse cases or discussion of minor themes? | | Manuscript (pgs. 9-14) |

### Supplemental Methods: Recruitment Procedures

The integrated health system consists of 11 hospitals and over 200 outpatient sites across 12 counties with 2.2 million system-wide outpatient registrations and an FQHC network that includes over 2 million patients across 70 organizations in 18 states. Within the integrated health system, recruitment methods included personalized, targeted emails sent to 42 prospective participants, a description of the project in the Primary Care Executive Council Research and Education Subcommittee Research & Education newsletter published six times between August 2022 and February 2023 and circulated to approximately 588 clinicians, and a presentation about the study at a health system-wide Heart Failure Quality Meeting. To recruit participants from FQHCs, we coordinated with an administrator of the FQHC network to send a recruitment email to 14 potentially eligible clinicians at FQHCs within the network. The recruitment email contained a hyperlink through which interested clinicians could schedule a 30-minute interview with a member of the study team via videoconference. For participants within the integrated health system, recruitment and data collection began in August 2022 and concluded in February 2023. For participants from FQHCs, recruitment and data collection began December 2022 and concluded March 2023. The initial target sample size was 30 cardiology and 30 primary care clinicians split across the different clinician types. Participants received a $100 stored value VISA card after completion of the interview.

### Supplemental Methods: Data Collection

Two team members (LPG and SEP), who have graduate-level training in qualitative data collection. The interviews were audio and video recorded with the files stored in a secure location. The interview guides, which are included in Tables 1,2,3, and 4, in the next section of the Supplement consisted of a structured set of questions, posed to each participant, and optional follow-up probes. Using CFIR, we developed questions to identify factors that impact the implementation of evidence-based care.^22,25^ We developed additional questions based on the Chronic Care Model, because this model includes specific elements of health systems associated with promoting high quality, evidence-based care.^26,27^ The guides were developed collaboratively by the research team and pilot tested during 2 interviews with cardiologists and 2 interviews with primary care physicians in the integrated health system. Following the pilot interviews, we refined the interview guides for clarity and prioritized sections in the event of time constraints. Demographic information, including age, ethnicity, race, and sex, was collected.

### Supplemental Methods: Analysis and Theoretical Framework

To report our results using the TPB-CFIR model, SEP first consolidated and refined the descriptions of the determinants and common themes prior to an additional review by FSA and LPG. Once we agreed on the description of the themes, SEP grouped them according to TPB construct and CFIR 2.0 domains.^25^ These groupings were then iteratively refined based on input from additional team members.

### Interview Guides

#### Table 2. Primary Care Clinician Interview Questions

1. **General**
   a. What is the main site where you practice?
   b. How many patients do you see with heart failure with reduced ejection fraction (HFrEF) in a typical month?
   c. How important is guideline directed medial therapy (GDMT) to the treatment of your patients with HFrEF?
2. **Care Coordination and Roles in Management of Medical Therapy**
   a. Whose role is it to manage the medical therapy of patients with HFrEF within your practice?
      i. How do physicians, NPs, PAs, RNs, and pharmacists collaborate on taking care of these patients?
      ii. When changes are made to a patient’s heart failure medication regimen, whose responsibility is it to follow up with patients with HFrEF about their medical therapy? What should this follow-up care entail?
      iii. Is this division of responsibility clearly understood by the members of the care team?
   b. For patients co-managed in cardiology practices, do you consider changes in heart failure meds when you see your patients in your clinic?
      i. If so, how do you communicate changes in medications and recommendations with cardiology clinicians who co-manage patients?
      ii. If not, what are the reasons you don’t consider changes in meds?
          1. *[Prompt for reasons]*
             a. Feelings about ownership over these medication changes
             b. Shared care with a cardiologist allows you to focus on other medical problems
             c. Thinks the cardiologist should be the only one making changes to heart failure medications if patient is under their care
   c. How do you approach the medical management of your patients with HFrEF who are not co-managed with cardiology?
3. **Prescribing Decision-Making**
   a. Do you feel comfortable prescribing and increasing the dose of the following classes of medications? ***[prompt for responses for both prescribing and increasing dose after each class]***
      i. Beta blockers
      ii. Angiotensin-converting enzyme inhibitors (ACE inhibitors) or angiotensin receptor blockers (ARBs)
      iii. Angiotensin receptor-neprilysin inhibitors (ARNIs), maybe also be referred to as Entresto
           1. Prompt-If not comfortable ask if they would be comfortable prescribing on low dose. Is discomfort around prescribing or increasing the dose or both?
      iv. Mineralocorticoid receptor antagonists (MRAs), such as spironolactone and eplerenone
      v. SGLT2 inhibitors (prescribing only)
   b. For patients not on all four classes of medications at maximally tolerated doses, other than contraindications, what factors affect whether you discuss and make recommendations to change their medication regimen?
      i. [Additional Prompts]
         1. What are some reasons that you would not recommend changes to a patient’s GDMT regimen?
         2. How do patient co-morbidities, frailty, and age affect your decision-making?
         3. If the patient is “doing well,” how does that affect your clinical decision making?”
         4. How do concerns around affordability and costs to patients affect your decision-making?
            a. [Prompt] Does medication class play a role in these concerns? For example, do these concerns come up more often for SGLT2 inhibitors and ARNIs because there aren’t generic options?
         5. How do concerns around other barriers to patient adherence to medication affect your decision-making?
         6. What role do you think “clinical inertia” plays in whether clinicians recommend a medication change during a clinic visit? Clinical inertia is the failure to make a guideline-recommended change in a patient’s medication regimen without a barrier or contraindication.
   c. Does optimization of GDMT take a backseat to other concerns when caring for your patients with HFrEF?
      i. If so, what other concerns are higher priority?
   d. Are there other resources available to help you manage complex patients with HFrEF?
      i. If so, which resources are most helpful?
      ii. If not, what resources or educational options would be most helpful (e.g., reminders, clinician education)?
4. **Self-Management Support**
   a. I asked earlier about how important GDMT is to the treatment of your patients with HFrEF. Do your patients feel similarly to you about how important GDMT is to the treatment of their heart failure?
   b. How receptive are your patients with HFrEF to making changes to their GDMT, such as the addition of a new medication or changes in dosing?
      i. What factors do you think affect their acceptance of changes in their medication regimen for HFrEF?
         1. What barriers prevent acceptance?
         2. What factors facilitates acceptance?
5. **Identifying Characteristics of an Ideal Health IT-Based Intervention**
   a. Feasibility as Solution
      i. Do you think any of the barriers to GDMT for patients with HFrEF could be addressed through a health IT-based intervention?
         1. If so, which ones and how?
         2. If none, why not?
   b. Experience with BPAs
      i. Are you familiar with best practice alerts (BPAs) within the EHR?
         1. If so, what has your experience been with them?
   c. Ideal BPA Design
      i. How would you design an alert to remind yourself to optimize prescription of GDMT for patients with HFrEF?
         1. Prompt for characteristics that influence alert fatigue or likelihood to pay attention to alert
         2. What information would be useful to you at the time of making prescribing decisions for these patients (e.g., current medications, recent lab data, link to guidelines)?
         3. Would the addition of an order set be helpful to facilitate adjustments to a patient’s medication?
   d. Integration into Workflow
      i. What should trigger these alerts?
      ii. How frequently should these alerts appear?
      iii. What is the next thing you would want to do after viewing the alert?
      iv. Are there any special considerations about your care setting or workflow that should influence how the alert is designed?
   e. Announcement of Changes/Rollout of New Workflow or Policy
      i. What factors should health system administrators consider when deciding whether to pilot or permanently adopt an alert like this?
      ii. If a new alert were being piloted or permanently adopted at your site, how would you like this information communicated to you?

**If Time Allows**

6. **Organization of Health Care**
   a. Does the health system, including clinician leaders, support adoption of evidence-based care and guideline recommendations?
   b. Are there any incentives, penalties, policies, or programs that make you more or less likely to prescribe GDMT for patients with HFrEF?
      i. If so, what are they? [Prompt for factors that are both internal and external to health system]
   c. What health system changes or improvement strategies may be effective in increasing prescription of GDMT for patients with HFrEF?
7. **Clinician Information Systems**
   a. Are you able to generate a list of patients with HFrEF under your care?
   b. Do you have access to information on your performance in providing GDMT to patients with HFrEF?
      i. Would you like access to your performance information?
         1. If not, why not?
         2. If so
            a. How would you like that information presented to you?
            b. How would you use this information?
      ii. Would you like access to information about the performance of your peers in prescribing GDMT to patients with HFrEF?
8. **Identifying Characteristics of an Ideal Health IT-Based Intervention**
   a. Announcement of Changes/Rollout of New Workflow or Policy
      i. Have you seen any changes rolled out in the past in ways that caused distrust or backlash from clinicians? If so, what do you think contributed to this response?
9. **Self Management Support**
   a. How confident are your patients in their ability to manage their medications for HFrEF?
      i. What skills are required for patients to be able to effectively manage their medications for HFrEF?
10. **Use of Guidelines**
    a. Do you have access to the latest evidence-based guidelines for care of patients with HFrEF?
       i. Where do you go to access these guidelines?
       ii. Is it convenient to access these guidelines?
           1. If not, what would make it more convenient to access them?
    b. Do you discuss GDMT and its importance with your patients when goal setting, action planning, or problem solving together?
    c. Do you reference guidelines in your clinical practice when seeing patients with HFrEF?
       i. If so, at what point in your workflow do you reference the guidelines?
       ii. If not, why not? (e.g., already know the info, not useful, not applicable to my patients, too hard to find, not enough time, patients not interested in changes)
    d. Are there other resources available to help you manage complex patients with HFrEF?
       i. If so, which resources are most helpful?
       ii. If not, what resources or educational options would be most helpful (e.g., reminders, clinician education)?

#### Table 3. Cardiology Clinician Interview Questions

1. **General**
   a. What is the main NM site where you practice?
   b. How many patients do you see with HFrEF in a typical month?
   c. How important is guideline directed medial therapy (GDMT) to the treatment of your patients with HFrEF?
2. **Care Coordination and Roles in Management of Medical Therapy**
   a. Whose role is it to manage the medical therapy of patients with HFrEF within your practice?
      i. How do physicians, NPs, PAs, RNs, and pharmacists collaborate on taking care of these patients?
      ii. When changes are made to a patient’s heart failure medication regimen, whose responsibility is it to follow up with patients with HFrEF about their medical therapy? What should this follow-up care entail?
      iii. Is this division of responsibility clearly understood by the members of the care team?
   b. For patients who also have a primary care clinician, does cardiology solely make changes related to heart failure medications, or is there active co-management with the primary care clinician?
      i. How do you communicate changes in medications and recommendations with primary care clinicians who co-manage patients?
   c. Are there other resources available to help you manage complex patients with HFrEF?
      i. If so, which resources are most helpful?
      ii. If not, what resources would be helpful (e.g., reminders, clinician education)?
3. **Prescribing Decision-Making**
   a. Do you feel comfortable prescribing and increasing the dose of the following classes of medications for HFrEF: *[prompt for responses for both prescribing and increasing dose after each class]*
      i. Beta blockers
      ii. Angiotensin-converting enzyme inhibitors (ACE inhibitors) or angiotensin receptor blockers (ARBs)
      iii. Angiotensin receptor-neprilysin inhibitors (ARNIs), maybe also be referred to as Entresto
      iv. Mineralocorticoid receptor antagonists (MRAs), such as spironolactone and eplerenone
      v. SGLT2 inhibitors (prescribing only)
   b. For patients not on all four classes of medications at maximally tolerated doses, other than contraindications, what factors affect whether you discuss and make recommendations to change their medication regimen?
      i. [Additional Prompts]
         1. How do patient co-morbidities, frailty, and age affect your decision-making?
         2. If the patient is “doing well,” how does that affect your clinical decision making?”
         3. How do concerns around affordability and costs to patients affect your decision-making?
            a. [Prompt] Does medication class play a role in these concerns? For example, do these concerns come up more often for SGLT2 inhibitors and ARNIs because there aren’t generic options?
         4. How do concerns around other barriers to patient adherence to medication affect your decision-making?
         5. What are some other reasons that you would not recommend changes to a patient’s GDMT regimen?
         6. What role do you think “clinical inertia” plays in whether clinicians recommend a medication change during a clinic visit? Clinical inertia is the failure to make a guideline-recommended change in a patient’s medication regimen without a barrier or contraindication.
   c. Does optimization of GDMT take a backseat to other concerns when caring for your patients with HFrEF?
      i. If so, what other concerns are higher priority?
4. **Self-Management Support**
   a. I asked earlier about how important GDMT is to the treatment of your patients with HFrEF. Do your patients feel similarly to you about how important GDMT is to the treatment of their heart failure?
   b. How receptive are your patients with HFrEF to making changes to their GDMT, such as the addition of a new medication or changes in dosing?
      i. What factors do you think affect their acceptance of changes in their medication regimen for HFrEF?
         1. What barriers prevent acceptance?
         2. What factors facilitates acceptance?
5. **Identifying Characteristics of an Ideal Health IT-Based Intervention**
   a. Feasibility as Solution
      i. Do you think any of the barriers to GDMT for patients with HFrEF could be addressed through a health IT-based intervention?
         1. If so, which ones and how?
         2. If none, why not?
   b. Experience with BPAs
      i. Are you familiar with best practice alerts (BPAs) within the EHR?
         1. If so, what has your experience been with them?
            a. [Prompt] What BPA characteristics influence alert fatigue or likelihood of paying attention to BPA?
   c. Ideal BPA Design
      i. How would you design an alert to remind yourself to optimize prescription of GDMT for patients with HFrEF?
         1. What information would be useful to you at the time of making prescribing decisions for these patients (e.g., current medications, recent lab data, link to guidelines)?
         2. Would the addition of an order set be helpful to facilitate adjustments to a patient’s medication?
   d. Integration into Workflow
      i. What should trigger these alerts?
      ii. How frequently should these alerts appear?
      iii. What is the next thing you would want to do after viewing the alert?
      iv. Are there any special considerations about your care setting or workflow that should influence how the alert is designed?
   e. Announcement of Changes/Rollout of New Workflow or Policy
      i. What factors should health system administrators consider when deciding whether to pilot or permanently adopt an alert like this?
      ii. If a new alert were being piloted or permanently adopted at your site, how would you like this information communicated to you?
      iii. Have you seen any changes rolled out in the past in ways that caused distrust or backlash from clinicians? If so, what do you think contributed to this response?

**If Time Allows:**

6. **Organization of Health Care**
a. Does the health system, including clinician leaders, support adoption of evidence-based care and guideline recommendations?
b. Are there any incentives, penalties, policies, or programs that make you more or less likely to prescribe GDMT for patients with HFrEF?
i. If so, what are they? [Prompt for factors that are both internal and external to health system]
c. What health system changes or improvement strategies may be effective in increasing prescription of GDMT for patients with HFrEF?
7. **Clinician Information Systems**
a. Are you able to generate a list of patients with HFrEF under your care?
b. Do you have access to information on your performance in providing GDMT to patients with HFrEF?
i. Would you like access to your performance information?
1. If not, why not?
2. If so
a. How would you like that information presented to you?
b. How would you use this information?
ii. Would you like access to information about the performance of your peers in prescribing GDMT to patients with HFrEF?
8. **Identifying Characteristics of an Ideal Health IT-Based Intervention**
a. Announcement of Changes/Rollout of New Workflow or Policy
i. Have you seen any changes rolled out in the past in ways that caused distrust or backlash from clinicians? If so, what do you think contributed to this response?
9. **Self-Management Support**
a. How confident are your patients in their ability to manage their medications for HFrEF?
i. What skills are required for patients to be able to effectively manage their medications for HFrEF?
10. **Use of Guidelines**
a. Do you have access to the latest evidence-based guidelines for care of patients with HFrEF?
i. Where do you go to access these guidelines?
ii. Is it convenient to access these guidelines?
1. If not, what would make it more convenient to access them?
b. Do you discuss GDMT and its importance with your patients when goal setting, action planning, or problem solving together?
c. Do you reference guidelines in your clinical practice when seeing patients with HFrEF?
i. If so, at what point in your workflow do you reference the guidelines and where do you access them?
ii. If not, why not? (e.g., already know the info, not useful, not applicable to my patients, too hard to find, not enough time, patients not interested in changes)

#### Table 4. Primary Care Pharmacist Interview Guide Questions

1. **General**
   a. What is the main NM site where you practice?
   b. How many patients do you see with HFrEF in a typical month?
   c. How important is guideline directed medial therapy (GDMT) to the treatment of your patients with HFrEF?
2. **Care Coordination and Roles in Management of Medical Therapy**
   a. Whose role is it to manage the medical therapy of patients with HFrEF within your practice?
      i. How do physicians, NPs, PAs, RNs, and pharmacists collaborate on taking care of these patients?
      ii. When changes are made to a patient’s heart failure medication regimen, whose responsibility is it to follow up with patients with HFrEF about their medical therapy? What should this follow-up care entail?
      iii. Is this division of responsibility clearly understood by the members of the care team?
   b. For patients who also have a cardiologist, is there active co-management with the cardiologist of changes to the patient’s heart failure medications?
      i. How do you communicate changes in medications or recommendations with cardiology clinicians who co-manage patients?
   c. Are there other resources available to help you manage complex patients with HFrEF?
      i. If so, which resources are most helpful?
      ii. If not, what resources would be helpful (e.g., reminders, clinician education)?
3. **Prescribing Decision-Making**
   a. Do you feel comfortable prescribing and increasing the dose of the following classes of medications for HFrEF: *[prompt for responses for both prescribing and increasing dose after each class]*
      i. Beta blockers
      ii. Angiotensin-converting enzyme inhibitors (ACE inhibitors) or angiotensin receptor blockers (ARBs)
      iii. Angiotensin receptor-neprilysin inhibitors (ARNIs), maybe also be referred to as Entresto
      iv. Mineralocorticoid receptor antagonists (MRAs), such as spironolactone and eplerenone
      v. SGLT2 inhibitors (prescribing only)
   b. What leads you to discuss a medication change with a patient’s primary clinician?
   c. For patients not on all four classes of medications at maximally tolerated doses, other than contraindications, what factors affect whether you change their medication regimen?
      i. [Additional Prompts]
         1. How do patient co-morbidities, frailty, and age affect your decision-making?
         2. If the patient is “doing well,” how does that affect your clinical decision making?”
         3. How do concerns around affordability and costs to patients affect your decision-making?
            a. [Prompt] Does medication class play a role in these concerns? For example, do these concerns come up more often for SGLT2 inhibitors and ARNIs because there aren’t generic options?
         4. How do concerns around other barriers to patient adherence to medication affect your decision-making?
         5. What are some other reasons that you would not recommend changes to a patient’s GDMT regimen?
         6. What role do you think “clinical inertia” plays in whether clinicians recommend a medication change during a clinic visit? Clinical inertia is the failure to make a guideline-recommended change in a patient’s medication regimen without a barrier or contraindication.
   d. Does optimization of GDMT take a backseat to other concerns when caring for your patients with HFrEF?
      i. If so, what other concerns are higher priority?
4. **Self-Management Support**
   a. I asked earlier about how important GDMT is to the treatment of your patients with HFrEF. Do your patients feel similarly to you about how important GDMT is to the treatment of their heart failure?
   b. How receptive are your patients with HFrEF to making changes to their GDMT, such as the addition of a new medication or changes in dosing?
      i. What factors do you think affect their acceptance of changes in their medication regimen for HFrEF?
         1. What barriers prevent acceptance?
         2. What factors facilitates acceptance?
5. **Identifying Characteristics of an Ideal Health IT-Based Intervention**
   a. Feasibility as Solution
      i. Do you think any of the barriers to GDMT for patients with HFrEF could be addressed through a health IT-based intervention?
         1. If so, which ones and how?
         2. If none, why not?
   b. Experience with BPAs
      i. Are you familiar with best practice alerts (BPAs) within the EHR?
         1. If so, what has your experience been with them?
            a. [Prompt] What BPA characteristics influence alert fatigue or likelihood of paying attention to BPA?
   c. Ideal BPA Design
      i. For HFrEF patients who can’t been seen in MAT clinic or aren’t manage primarily in cardiology, how would you design an alert to encourage their clinicians to optimize prescription of GDMT?
         1. What information would be useful to clinicians at the time of making prescribing decisions for these patients (e.g., current medications, recent lab data, link to guidelines)?
         2. Would the addition of an order set be helpful to facilitate adjustments to a patient’s medication?
   d. Integration into Workflow
      i. What should trigger these alerts?
      ii. How frequently should these alerts appear?
      iii. What is the next thing you would want to do after viewing the alert?
      iv. Are there any special considerations about your care setting or workflow that should influence how the alert is designed?
   e. Announcement of Changes/Rollout of New Workflow or Policy
      i. What factors should health system administrators consider when deciding whether to pilot or permanently adopt an a health IT based intervention like this?
      ii. If a new health IT based intervention were being piloted or permanently adopted at your site, how would you like this information communicated to you?
      iii. Have you seen any changes rolled out in the past in ways that caused distrust or backlash from clinicians? If so, what do you think contributed to this response?

**If Time Allows:**

6. **Organization of Health Care**
a) Does the health system, including clinician leaders, support adoption of evidence-based care and guideline recommendations?
b) What health system changes or improvement strategies may be effective in increasing prescription of GDMT for patients with HFrEF?
7. **Self-Management Support**
   a) How confident are your patients in their ability to manage their medications for HFrEF?
   b) What skills are required for patients to be able to effectively manage their medications for HFrEF?

#### Table 5. Cardiology Pharmacist Interview Guide Questions

1. **General**
   a. What is the main NM site where you practice?
   b. How many patients do you see with HFrEF in a typical month?
   c. How important is guideline directed medial therapy (GDMT) to the treatment of your patients with HFrEF?
2. **Care Coordination and Roles in Management of Medical Therapy**
   a. Whose role is it to manage the medical therapy of patients with HFrEF within your practice?
      i. How do physicians, NPs, PAs, RNs, and pharmacists collaborate on taking care of these patients?
      ii. When changes are made to a patient’s heart failure medication regimen, whose responsibility is it to follow up with patients with HFrEF about their medical therapy? What should this follow-up care entail?
      iii. Is this division of responsibility clearly understood by the members of the care team?
   b. For patients who also have a primary care clinician, does cardiology solely make changes related to heart failure medications, or is there active co-management with the primary care clinician?
      i. How do you communicate changes in medications and recommendations with primary care clinicians who co-manage patients?
   c. Are there other resources available to help you manage complex patients with HFrEF?
      i. If so, which resources are most helpful?
      ii. If not, what resources would be helpful (e.g., reminders, clinician education)?
3. **Prescribing Decision-Making**
   a. Do you feel comfortable prescribing and increasing the dose of the following classes of medications for HFrEF: *[prompt for responses for both prescribing and increasing dose after each class]*
      i. Beta blockers
      ii. Angiotensin-converting enzyme inhibitors (ACE inhibitors) or angiotensin receptor blockers (ARBs)
      iii. Angiotensin receptor-neprilysin inhibitors (ARNIs), maybe also be referred to as Entresto
      iv. Mineralocorticoid receptor antagonists (MRAs), such as spironolactone and eplerenone
      v. SGLT2 inhibitors (prescribing only)
   b. What leads you to discuss a medication change with a patient’s primary clinician?
   c. For patients not on all four classes of medications at maximally tolerated doses, other than contraindications, what factors affect whether you change their medication regimen?
      i. [Additional Prompts]
         1. How do patient co-morbidities, frailty, and age affect your decision-making?
         2. If the patient is “doing well,” how does that affect your clinical decision making?”
         3. How do concerns around affordability and costs to patients affect your decision-making?
            a. [Prompt] Does medication class play a role in these concerns? For example, do these concerns come up more often for SGLT2 inhibitors and ARNIs because there aren’t generic options?
         4. How do concerns around other barriers to patient adherence to medication affect your decision-making?
         5. What are some other reasons that you would not recommend changes to a patient’s GDMT regimen?
         6. What role do you think “clinical inertia” plays in whether clinicians recommend a medication change during a clinic visit? Clinical inertia is the failure to make a guideline-recommended change in a patient’s medication regimen without a barrier or contraindication.
   d. Does optimization of GDMT take a backseat to other concerns when caring for your patients with HFrEF?
      i. If so, what other concerns are higher priority?
4. **Self-Management Support**
   a. I asked earlier about how important GDMT is to the treatment of your patients with HFrEF. Do your patients feel similarly to you about how important GDMT is to the treatment of their heart failure?
   b. How receptive are your patients with HFrEF to making changes to their GDMT, such as the addition of a new medication or changes in dosing?
      i. What factors do you think affect their acceptance of changes in their medication regimen for HFrEF?
         1. What barriers prevent acceptance?
         2. What factors facilitates acceptance?
5. **Identifying Characteristics of an Ideal Health IT-Based Intervention**
   a. Feasibility as Solution
      i. Do you think any of the barriers to GDMT for patients with HFrEF could be addressed through a health IT-based intervention?
         1. If so, which ones and how?
         2. If none, why not?
   b. Experience with BPAs
      i. Are you familiar with best practice alerts (BPAs) within the EHR?
         1. If so, what has your experience been with them?
            a. [Prompt] What BPA characteristics influence alert fatigue or likelihood of paying attention to BPA?
   c. Ideal BPA Design
      i. For HFrEF patients who can’t been seen in MAT clinic or aren’t manage primarily in cardiology, how would you design an alert to encourage their clinicians to optimize prescription of GDMT?
         1. What information would be useful to clinicians at the time of making prescribing decisions for these patients (e.g., current medications, recent lab data, link to guidelines)?
         2. Would the addition of an order set be helpful to facilitate adjustments to a patient’s medication?
   d. Integration into Workflow
      i. What should trigger these alerts?
      ii. How frequently should these alerts appear?
      iii. What is the next thing you would want to do after viewing the alert?
      iv. Are there any special considerations about your care setting or workflow that should influence how the alert is designed?
   e. Announcement of Changes/Rollout of New Workflow or Policy
      i. What factors should health system administrators consider when deciding whether to pilot or permanently adopt a health IT based intervention like this?
      ii. If a new health IT based intervention were being piloted or permanently adopted at your site, how would you like this information communicated to you?
      iii. Have you seen any changes rolled out in the past in ways that caused distrust or backlash from clinicians? If so, what do you think contributed to this response?

**If Time Allows:**

8. **Organization of Health Care**
c) Does the health system, including clinician leaders, support adoption of evidence-based care and guideline recommendations?
d) What health system changes or improvement strategies may be effective in increasing prescription of GDMT for patients with HFrEF?
9. **Self-Management Support**
   a) How confident are your patients in their ability to manage their medications for HFrEF?
   b) What skills are required for patients to be able to effectively manage their medications for HFrEF?

### Supplemental Methods: Coding Process Detailed Overview

Due to the addition of and refinement of codes after the initial round of coding, LPG and SEP recoded the initial 20% of the integrated health systems transcripts and FQHC transcripts and met again to confer on coding discrepancies. The additions and the changes to the code definitions were logged to support the potential replication of the coding process. This second round yielded good inter-rater reliability (α: 0.81).^30^ SEP coded the remaining 21 transcripts from the integrated health system participants and 6 transcripts for FQHC participants.

### Analytic Memo

#### Table 6. Analytic Memo Questions

1) What are the key clinician-level determinants to achieving target dosing of GDMT for patients with HFrEF?
2) What are the key patient-level determinants to achieving target dosing of GDMT for patients with HFrEF?
3) Where do these key determinants fall within the elements of a health care system defined in the Chronic Care Model?**

- The community
- The health system
- Self-management support
- Delivery system design
- Decision Support
- Clinical information systems

4) What are ideal components of an intervention that could help drive sustained intensification of GDMT?

- By what mechanism would these components of the intervention help produce productive interactions between an informed, activated patient and a prepared practice team?

5) Are there any major differences in responses from primary care clinicians vs. cardiology clinicians?
6) Are there any major differences in responses from primary care clinicians from NM vs. AllianceChicago?

** We originally planned to present our findings within the context of the Chronic Care Model, but following the discussion of the analytic memos, the decision was made to use an integrated model, consisting of the Theory of Planned Behavior and Consolidated Framework for Implimentation Research (CFIR) 2.0 instead.
